# Statistical, Multi-scale and Attention-based Layer Pooling of Wav2Vec-2 Speech Embeddings for Parkinson’s Disease Detection

**DOI:** 10.1101/2025.09.13.25335694

**Authors:** Ondrej Klempir, Juliana Grand Mullerova, Radim Krupicka

## Abstract

Self-supervised pre-trained speech models such as wav2vec 2.0 provide rich frame-level embeddings that are increasingly used for clinical voice screening, including Parkinson’s disease (PD). Optimally aggregating the frame-level embeddings is a significant task, with the underexplored question of how to best aggregate the frame-level embeddings into fixed-length utterance descriptors for the downstream task of binary classification (healthy controls (HC) vs. PD). To address this, our study compared three wav2vec 2.0 variants, the base model and two fine-tuned variants (one adapted for dysarthric corpora), across multiple layer depths, ten statistical aggregation functions (e.g., mean, median, quantiles), and two proposed advanced schemes (attention-based and multi-scale average pooling). We used the MDVR-KCL as a read-speech corpus (16 PD, 21 HC). Contrary to the expectation that sophisticated pooling would help, statistical aggregations such as mean pooling consistently provided better performance and robustness. Early representations (pre-Transformer and 1^st^ Transformer block) were often the most informative, and mean aggregation produced relatively high, low-variance scores across models and depths. Attention and multi-scale pooling did not yield consistent gains. Moreover, wav2vec-based embeddings outperformed traditional acoustic baselines. Applying the supervised feature selection (ANOVA F-value) further improved performance, i.e. conservative selection (12 features) achieved a mean balanced accuracy of 0.87 and precision of 0.92, with top configurations exceeding a 0.93 balanced accuracy and had a precision of 1.0. The findings empirically support the continued use of mean pooling as a viable strategy for temporal aggregation of latent features for PD wav2vec-based detection.

## 1. Introduction

Since the early 2010s, deep neural networks (NNs) have reshaped machine learning, displacing the earlier Gaussian mixture model–Hidden Markov model (GMM–HMM) pipelines in speech recognition and delivering substantial accuracy gains across vision, speech and other domains [1, 2, 3]. In the 2020’s a second revolution emerged, i.e. large-scale pre-training and self-supervised learning driven by Transformer architectures and large pre-trained models (for example, GPT-3), combined with self-supervised speech models such as wav2vec 2.0 [4]. These developments produced another disruptive leap in performance and greatly reduced the need for labelled data [4, 5, 6]. Progress in large pre-trained and self-supervised models opened new opportunities to extract subtle early signals of neurological disease from everyday data such as speech. Artificial Intelligence (AI)-powered early disease detection has marked a bold shift from reactive treatment to proactive prevention, catching silent threats before they are manifest. Smart scanning technologies and digital biomarkers converged to form a powerful fusion for early Parkinson’s disease (PD) detection [7].

PD is a common age-related neurodegenerative disorder whose core motor signs include bradykinesia, resting tremor and rigidity, and whose prevalence rises as the population ages [8]. A long prodromal phase, often including non-motor features such as hyposmia, constipation and REM-sleep (rapid eye movement) behaviour disorder, can precede the classical motor syndrome by many years; this makes early, scalable biomarkers particularly valuable [9, 10]. Because disease-modifying therapies have not yet been proven, an urgent need remains for sensitive methods that identify diseases earlier and that can track progression for both clinical trials and patient care [11, 12]. Voice and speech are commonly affected in PD, and these changes can appear early. Recent reviews indicate that speech and language signals are promising, non-invasive and scalable biomarkers for detection and monitoring when combined with modern machine learning approaches [13, 14, 15, 16].

Many practical machine-learning tasks in medicine suffer from class imbalance, high-dimensional or noisy features, heterogeneous recording conditions and datasets that are either very small or extremely large; these problems can substantially degrade model performance if they are not addressed [17]. These challenges are especially acute in PD speech research because clinically verified PD speech corpora are relatively small, vary by language and recording protocol, and often contain heterogeneous tasks and labels, which limits the effectiveness of purely supervised approaches [13, 18, 19]. As a result, pre-trained and self-supervised models that learn rich audio representations from a large amount of unlabelled speech, for example wav2vec 2.0 [4], have been widely adopted [19, 20, 21]. These models have reduced dependence on labelled data, improved robustness across recording conditions and have produced strong results on downstream PD detection tasks, therefore they are essential to consider when designing practical systems.

Classical machine-learning classifiers such as k-nearest neighbours, support vector machines and simple feed-forward networks remain relevant to the healthy controls (HC) versus PD detection tasks [22, 23, 24]. However, many of these programs operated on preprocessed, hand-engineered feature sets rather than on end-to-end raw signal modelling, which limits their ability to exploit the rich representations learned by pretrained audio models. At the same time, there is an observable shift toward quantum-inspired machine-learning classifiers for multimodal PD screening [25].

Recent research showed that speech foundation models, such as wav2vec 2.0/XLSR and other large pre-trained encoders, improved PD detection when used as either frozen feature extractors or when adapted by fine-tuning. For example, a recent cross-study using foundation models plus speech enhancement reported strong in-lab gains on PC-GITA but also highlighted fragility under cross-condition evaluations [26]. A study directly comparing layer selection, that is, choosing embeddings from particular transformer layers, versus model adaptation found that careful layer-wise probing can be competitive with full fine-tuning [27]. A multiple sclerosis layer-wise study (XLSR wav2vec 2.0 fine-tuned on Hungarian) found that the lowest one-third of the 24 fine-tuned layers proved to be the most suitable for feature extraction [28].

A neglected bottleneck is how frame-level embeddings are aggregated into fixed-length utterance representations. In other words, how to handle utterances of different durations. Vetrab et al. indicated that aggregating frame-level embeddings is a task that remains significant [29]. The choice of which aggregation method to use is often overlooked, and simple functions such as mean and standard deviation have been commonly used without solid experimental justification, especially for PD detection using speech and voice. Beyond wav2vec, the authors have applied mean and max pooling as well as majority voting on Transformer-based systems [30]. PD wav2vec-based studies have most often used simple mean pooling [27, 31, 32]. Evidence from related tasks and from recent research on attention-based feature fusion, multi-scale pooling and recurrent decoders such as gated recurrent unit (GRU)-based aggregation suggested that weighted approaches or multi-scale pooling can improve robustness to variable utterance duration and can emphasize diagnostically relevant frames [33, 34, 35].

The aforementioned findings motivated our experiments, which (A) compared layer selection with different fine-tuning wav2vec 2.0 recipes, and (B) evaluated mean pooling against alternative statistical and advanced pooling strategies, including attention and multi-scale average pooling, to test whether the prevailing mean-pool choice in PD research is empirically justified.

The main contributions of this work are as follows:

◯ We evaluated three wav2vec2 variants for PD detection, including quantifying the base model and the effect of fine-tuning using a dysarthric speech corpus.
◯ We compared the commonly used mean aggregation against other alternative statistical aggregation strategies to assess whether mean pooling is justified for PD detection.
◯ We proposed advanced wav2vec2 layer-pooling methods, specifically attention-based pooling and multi-scale average pooling, and evaluated their impact on classification performance.
◯ We investigated feature selection on wav2vec-derived embeddings and their impact on classification performance.

## 2. Methods

In Fig. 1, the proposed wav2vec-based methodology is illustrated.

**Fig. 1.**
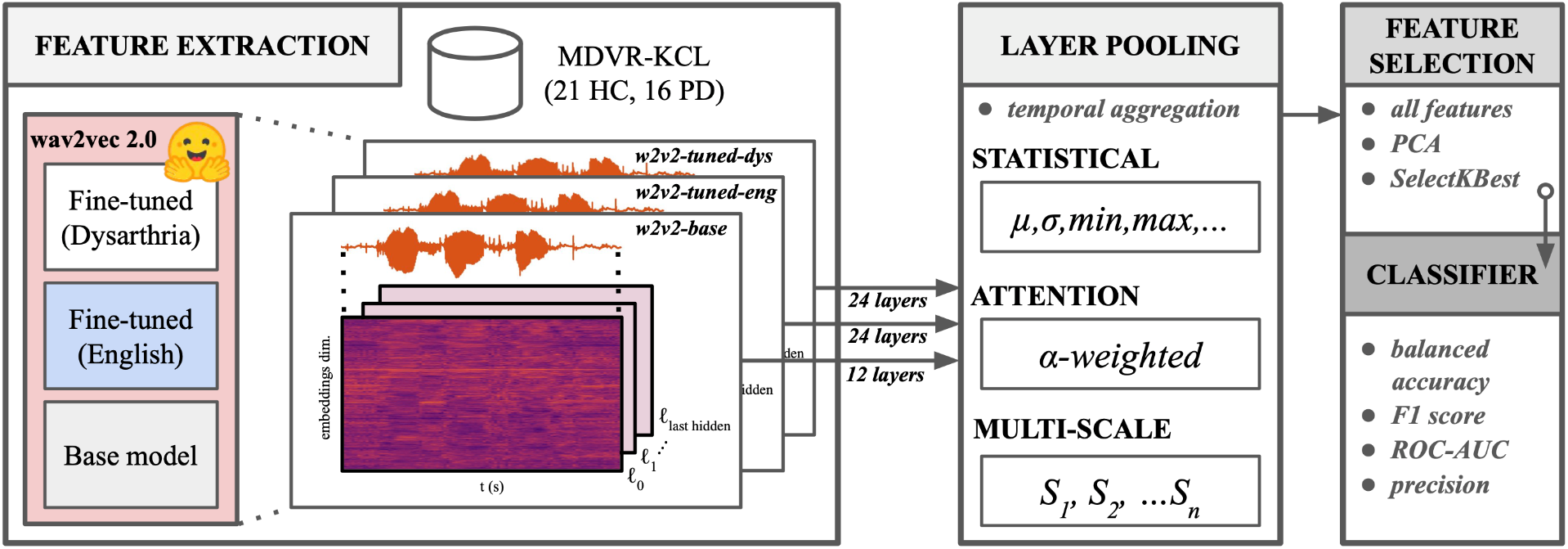
An overview of the methodology for Parkinson’s disease classification using wav2vec 2.0 embeddings. Different layer pooling strategies (statistical, attention-based, and multi-scale) were evaluated using three pre-trained models with feature selection techniques applied prior to supervised classification. The waveform (orange) displays a two-second speech segment with visibly different amplitude patterns and temporal organization. Below, the hierarchically clustered 768-dimensional embeddings from a selected layer show how the neural model w2v2-base represents this speech segment, with similar embedding dimensions grouped together vertically (hierarchically clustered). Additionally, a set of traditional acoustic features was extracted for comparative analysis (not shown).

### 2.1. Speech Corpus

The English MDVR-KCL dataset [36] comprised of a set of voice recordings collected in a clinical setting at King’s College London Hospital. Using a Moto G4 smartphone and a custom recording app, uncompressed .wav audio at *f*_*s*_ = 44. 1 *kHz* sample rate captured participants reading fixed text passages “The North Wind and the Sun” and a technical geography snippet, as well as engaging in spontaneous dialogue. Each recording is labelled with health status (PD = 1, HC = 0), Hoehn & Yahr scale, and UPDRS-II part 5 and UPDRS-III part 18 scores. In this study, we focused exclusively on fixed read text recordings, retaining full passages without editing. The corpus included 16 PD and 21 HC subjects. A typical waveform recording for both groups is visualized in Fig. 2.

**Fig. 2.**
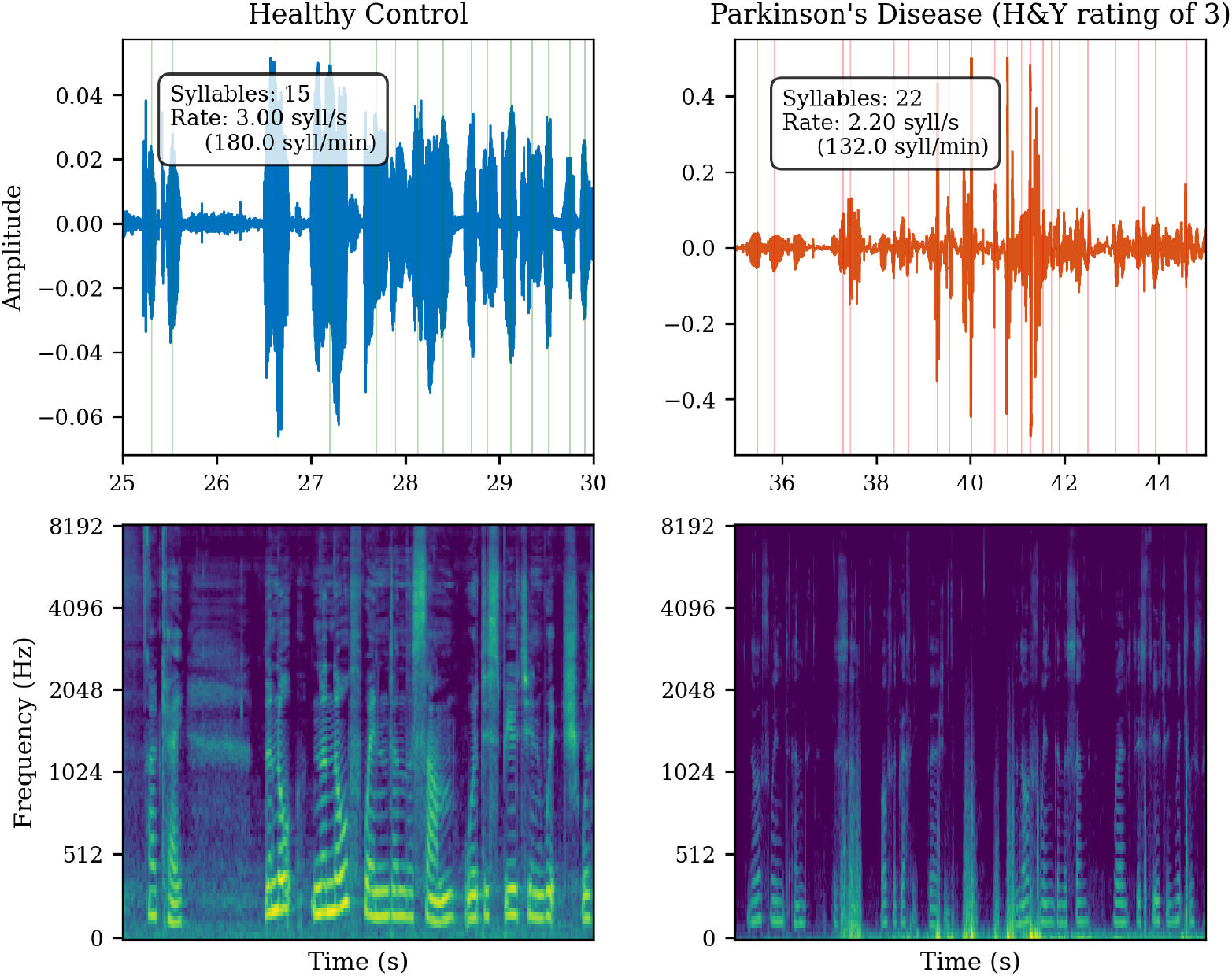
A visual inspection of speech characteristics between HC and PD subjects. The top panels display waveforms with estimated syllable boundaries (vertical lines) and calculated speech rate statistics, while the bottom panels show corresponding spectrograms (frequency limited to 8000 Hz). Most speech energy was concentrated below 5000 Hz. Vowel formants typically range from 300-3500 Hz, while consonant information extends to about 8000 Hz. HC speech (5 seconds duration; left) and PD speech (10 seconds duration; right) demonstrated waveform and spectrogram differences in energy distribution for the fixed reading passage “The North Wind and the Sun were disputing which was the stronger”.

### 2.2. Audio Pre-processing

Let **x** = [*x*_0_, …, *x*_*N-*1_ ] denote the time-domain signal. All recordings were resampled to a uniform sampling rate of *f*_*s*_ = 16 *kHz* using high-quality sinc interpolation. The signals were then peak-normalized to 95% of the full-scale range, such that 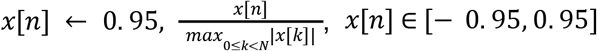.

### 2.3. Traditional Signal-processing Features

#### 2.3.1. Audio Recording Duration Features

To evaluate whether the total duration of a signal can predict PD status, we extracted both the total speech duration and loud-speech duration for each recording. The total duration in seconds is computed as 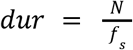, where N is the number of samples. The loud-speech duration (in seconds) was estimated using a sliding-window approach. A frame length of w = 0.025 seconds (400 samples at 16 kHz) and hop size of h = 0.010 seconds (160 samples) was applied.

The short-term frame energy for frame index *m* was given by 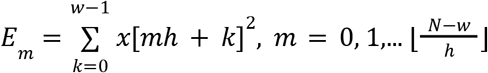 A median-based threshold *θ* = *median*(*E*_*m*_) was used to distinguish voiced (loud) frames, where a frame was classified as *loud* if *E* > θ. The total loud-speech duration was then 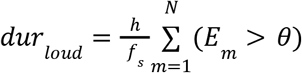 Taken together, these two measures formed a two-dimensional feature vector **b**_*dur*_ = [*dur, dur*_*loud*_ ] ∈ *R*^2^ .

#### 2.3.2. Acoustic Descriptors

A set of baseline acoustic descriptors was extracted from each audio recording (Table 1). Where descriptors produced a temporal trajectory *z*_*t*_, the following set of statistical functionals was consistently applied:

**Table 1.**
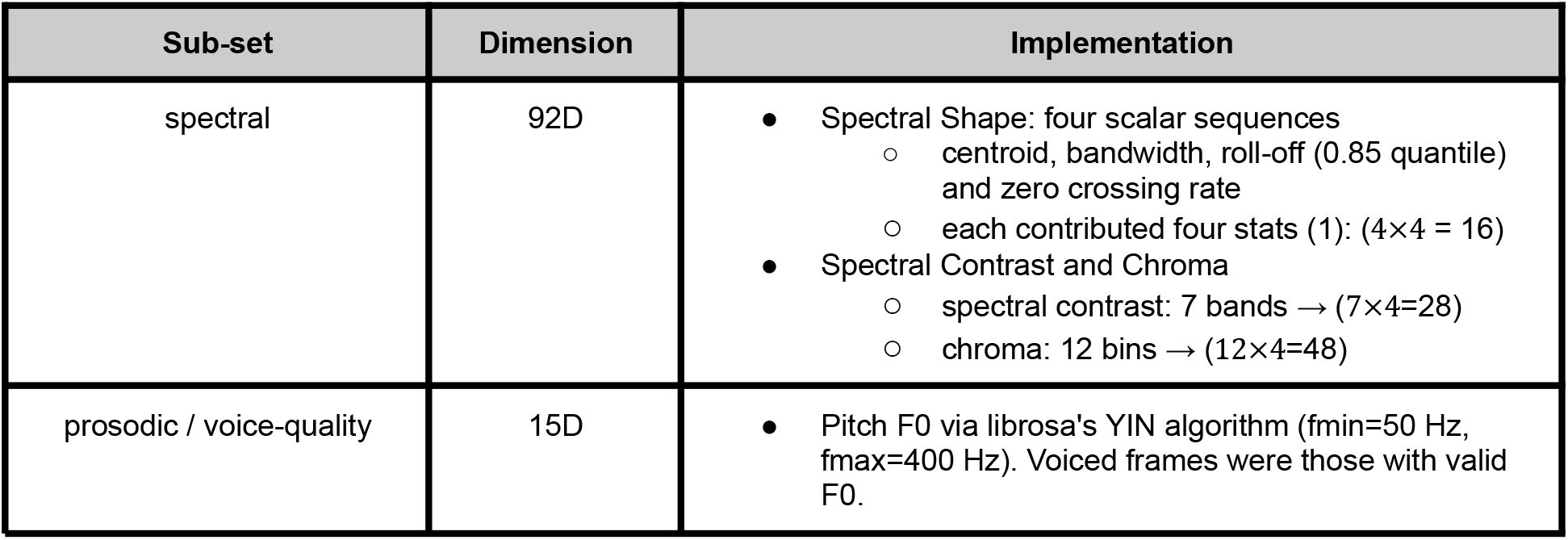
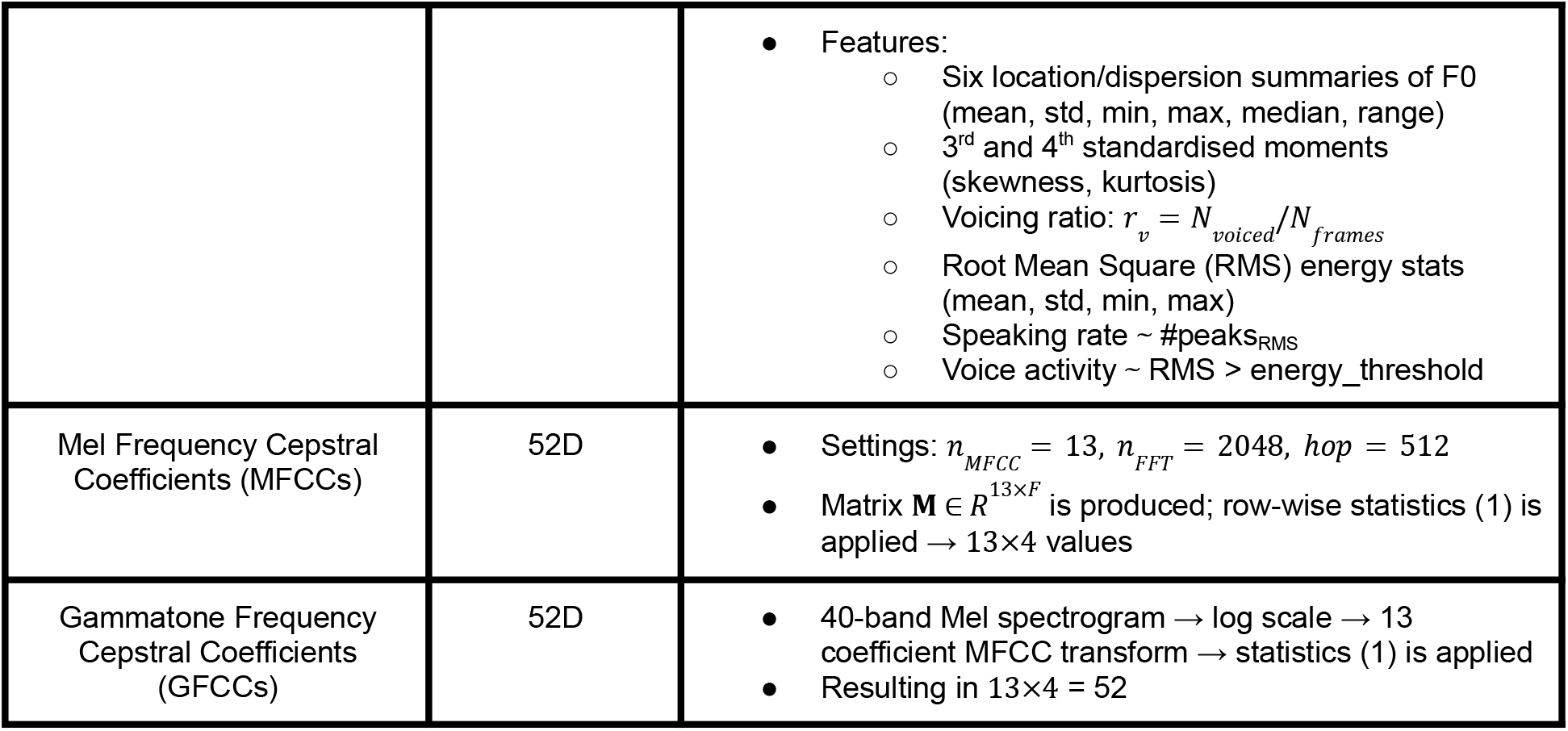
List of calculated low-level descriptors.

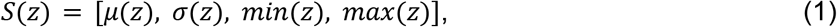

applying these summary statistics to all applicable descriptors yielded a fixed-length feature representation for each recording. The resulting baseline acoustic features are denoted as **b** _*acoustic*_ ∈ *R*^211^ .

### 2.4. Wav2Vec-2.0 Embeddings

#### 2.4.1. Pre-trained Self-supervised Models

A pre-trained model is typically a large NN trained in advance on a substantial volume of data, enabling it to learn general-purpose representations. In self-supervised learning, the model learns to predict parts of the input from other observed parts, without requiring manual labels. For speech models such as Wav2Vec 2.0, this involves masking portions of the audio signal in the latent space and training the network to reconstruct them from surrounding context, capturing both phonetic and prosodic characteristics.

We used Wav2Vec 2.0 embeddings as fixed-dimensional feature generators (Table 2), converting variable-length recordings into classification-ready vectors. All model weights were kept frozen (no further fine-tuning) to preserve the original learned representations. All audio was normalized to a 16 kHz sampling rate before embedding extraction. We employed three architectures:

**Table 2.**
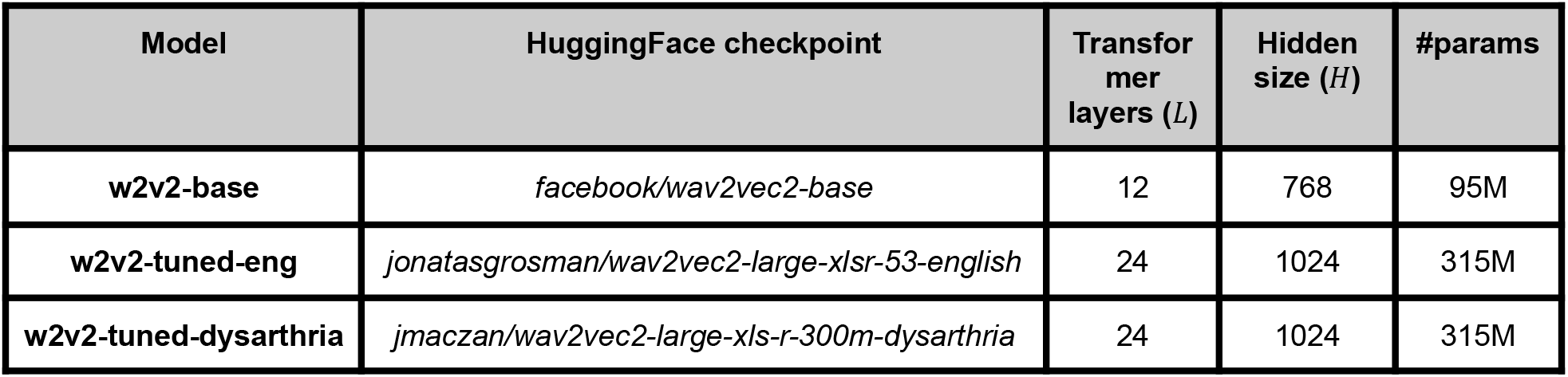
List of pre-trained Wav2Vec 2.0 models (all released under the Apache License 2.0).

1. ***wav2vec2-base*** [37]: Pre-trained on 16 kHz speech from the Librispeech corpus (∼960 h of read English audiobooks).
2. ***wav2vec2-large-xlsr-53-english*** [38]: Based on the multilingual wav2vec2-large-xlsr-53 model, pre-trained on 56,000 h of speech across 53 languages, then fine-tuned on the English subset of Common Voice 6.1. The dataset includes English speech (e.g., sentences such as “She’ll be all right.”), which we hypothesized might better capture accent-specific prosodic and articulatory cues relevant for detecting dysarthria in the MDVR-KCL PD corpus.
3. ***wav2vec2-large-xls-r-300m-dysarthria*** [39]: Derived from the wav2vec2-large-xls-r-300m model and further fine-tuned on two dysarthric speech corpora: TORGO [40] and UASpeech [41]. While neither corpus specifically targets PD, both contain audio recordings with dysarthric characteristics.

#### 2.4.2. Layer Selection and Feature Generator

For each pre-trained model, we considered a subset of layer outputs to balance representational diversity and computational efficiency. Specifically, we sampled at least layer indices *ℓ* ∈ {0, 1, ⌊*L*/2⌋, *L* − 1} where *L* is the total number of Transformer layers. By doing so, we captured representations from early, mid, and advanced stages of the model without needing to exhaustively evaluate every layer:

- *ℓ* = 0: output of the convolutional feature encoder (pre-Transformer)
- *ℓ* = 1: output of the first Transformer block, reflecting early contextualization of acoustic features
- *ℓ* = ⌊*L*/2⌋: output of the central Transformer block, representing mid-level abstractions
- *ℓ* = *L* − 1: output of the second-to-last Transformer block, capturing high-level contextual features

This layered design supported extracting features from different depths and offered representations that varied from raw acoustic to high-level contextual information. For each audio chunk, the model output hidden states across layers. Given the hidden activation tensor from layer *ℓ* for a single utterance:

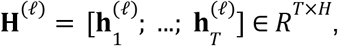

where *T* is the sequence length (number of frames) and *H* is the hidden dimensionality.

To handle long recordings, we applied chunking, i.e. audio longer than *N* > 160,000 (≈ 10 s at 16 kHz) was split into non-overlapping segments of ≤ 10 s. For each chunk, hidden states were extracted independently and then concatenated along the temporal dimension prior to downstream aggregation. Thus, a long waveform is partitioned into equal-length chunks, embeddings are computed per chunk, and the resulting hidden sequences are merged into a continuous representation before feature aggregation.

#### 2.4.3. Statistical Pooling

For each feature axis *d* = 1 … *H*, ten temporal aggregation (mean, std, max, min, median, q25, q75, iqr, skewness, kurtosis) functions ϕ_*j*_ were defined (Table 3).

**Table 3.** List of statistical temporal aggregation functions.

| $j$ | Operator $\phi_j$ | Formula |
| --- | --- | --- |
| 1 | mean | $\mu_d = \frac{1}{T} \sum_{t=1}^T H_{t,d}^{(\ell)}$ |
| 2 | standard deviation | $\sigma_d = \sqrt{\frac{1}{T} \sum_t \left( H_{t,d}^{(\ell)} - \mu_d \right)^2}$ |
| 3 | maximum | $\max_t \left( H_{t,d}^{(\ell)} \right)$ |
| 4 | minimum | $\min_t \left( H_{t,d}^{(\ell)} \right)$ |
| 5 | median | $\text{median}_t \left( H_{t,d}^{(\ell)} \right)$ |
| 6 | 25<sup>th</sup> percentile | $q_{25,d}$ |
| 7 | 75<sup>th</sup> percentile | $q_{75,d}$ |
| 8 | inter-quartile range | $IQR_d = q_{75,d} - q_{25,d}$ |
| 9 | skewness | *scipy.stats.skew* [42] |
| 10 | kurtosis | *scipy.stats.kurtosis* [43] |

Each ϕ_*j*_ returns a vector in *R*^*H*^ ; concatenation is not performed, i.e. the resulting vectors were treated as independent feature types (768 or 1024 dim. size).

#### 2.4.4. Attention-based Pooling

To emphasize salient frames without introducing any trainable parameters, we employed a non-learning attention pooling mechanism. This approach assigns higher weight to frames based on the magnitude (L1 norm) of their feature vectors and treats high-energy frames as more informative. This design de-emphasizes long silences, as frames with near-zero magnitude yield near-zero weights after softmax normalization.

For each frame-level vector **h** ∈ *R*^*H*^, we compute its salience score as the L1 norm, 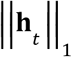 . The attention weight α_*t*_ is then determined via α_*t*_ 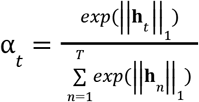. The aggregated embedding is the weighted sum 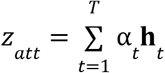. This attention pooling method combined local importance (via frame salience) with a global normalization (768 or 1024 dim. size).

#### 2.4.5. Multi-scale Average Pooling

To capture temporal patterns at multiple resolutions, we employed multi-scale average pooling, which aggregated frame-level representations over several window sizes and merged them into a single fixed-length descriptor. Unlike simple mean pooling, this approach summarizes both fine-grained and long-range information, without introducing additional learnable parameters. Intuitively, small kernel sizes (*k*) capture fine phonetic detail, while larger kernels emphasize broader prosodic or rhythmic trends. Averaging across scales reduces sensitivity to segmentation boundaries and might increase robustness against long silences, since larger kernels smooth out silent regions. If the input sequence was shorter than the smallest non-trivial kernel, the procedure defaulted to global mean pooling to ensure numerical stability.

##### Algorithm 1

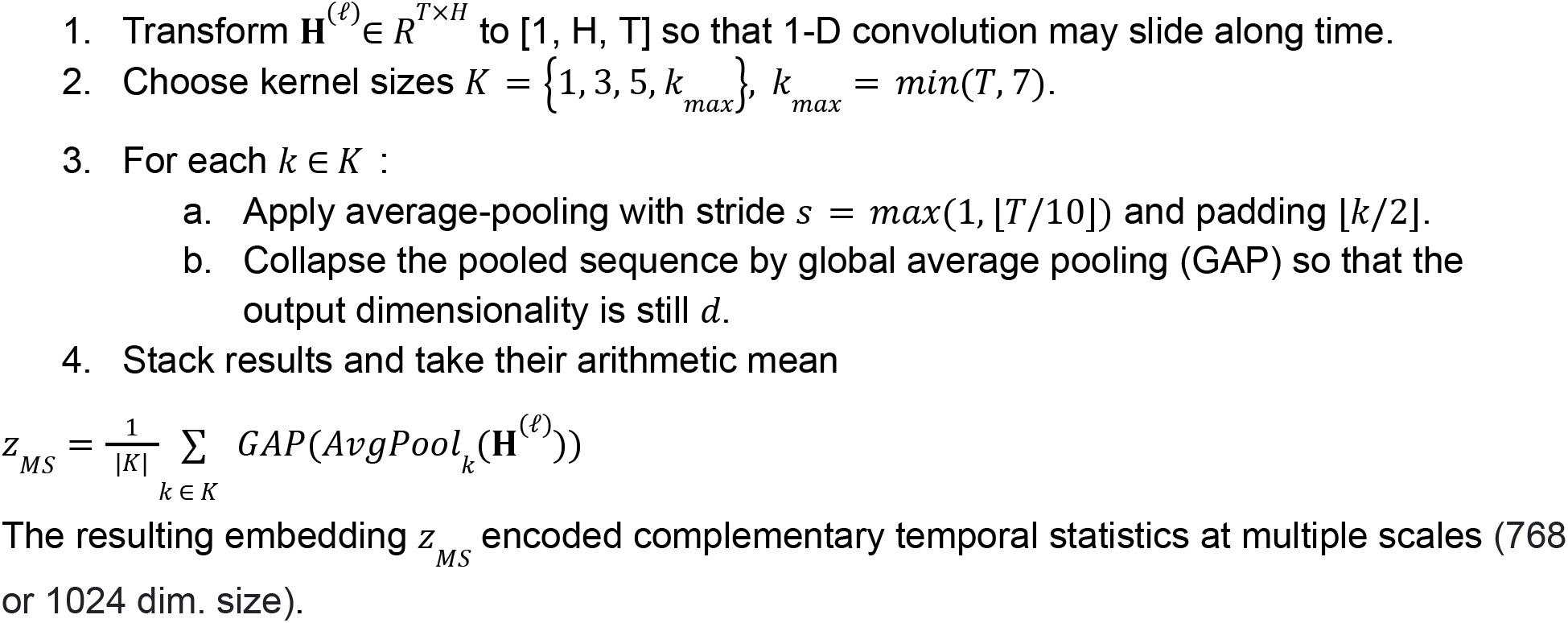

### 2.5. Supervised Classification

#### 2.5.1. Learners

The prediction task was formulated as a binary classification problem, distinguishing between HC (label = 0) and patients with PD (label = 1). The unit of classification was the utterance, i.e., each individual recording received an independent label prediction. Model performance was estimated using 5-fold stratified cross-validation. As the primary learner, we employed a Random Forest (RF) classifier with 100 trees and fixed random seed (*n*_estimators_=100, random_state=42). The choice of RF was motivated by its robustness to noisy or redundant features, its built-in feature selection via split criteria, and its ability to handle imbalanced datasets without requiring extensive hyperparameter tuning.

For hand-crafted acoustic baselines, features were standardised within each training fold using z-normalisation, 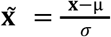, where *μ* and *σ* denote the mean and standard deviation. In contrast, when training tree-based models on wav2vec-derived embeddings, we used raw, non-normalised vectors.

#### 2.5.2. Feature Selection

To reduce dimensionality and focus on the most informative predictors, we considered two complementary feature selection strategies. First, the ANOVA F-value with the SelectKBest method was applied [44], ranking features according to their statistical relevance with respect to binary labels and retaining the top *k*. Second, we applied Principal Component Analysis (PCA), projecting the original feature space into an orthogonal basis and keeping *min*(*k, N* − 1, *p*) principal components, where *N* is the number of samples and *p* is the original feature dimensionality.

We defined two operating modes for feature selection:

- Conservative mode, yielding 12 top features,
- Moderate mode, yielding 20 top features.

#### 2.5.3. Evaluation Metrics

Model performance was assessed using multiple complementary evaluation metrics to capture different aspects of predictive quality and to account for class imbalance. The primary evaluation metric was ***Balanced Accuracy*** (BA), defined as the arithmetic mean of sensitivity (true positive rate) and specificity (true negative rate):

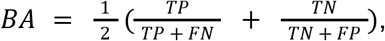

Where *TP, TN, FP, FN*, denote true positives, true negatives, false positives, and false negatives, respectively.

In addition, the following metrics were reported:

- ***F1 Score:*** harmonic mean of precision and recall, capturing the trade-off between detecting PD and avoiding false alarms.
- ***ROC-AUC:*** area under the receiver operating characteristic curve, quantifying the ability to discriminate between classes across thresholds.
- ***Precision:*** proportion of predicted PD cases that were correctly classified, reflecting the reliability of positive predictions.

### 2.6. Computational Environment

All experiments were conducted on an Amazon Web Services (AWS)-EC2 g4dn.xlarge instance equipped with a 16 GB NVIDIA T4 Tensor Core GPU. The computational workflow was managed within a Databricks environment, using Python 3.10 with the following key libraries: NumPy 1.24 for numerical computation, PyTorch 2.0 for deep learning model implementation, Transformers 4.32 for pre-trained model handling, and Librosa 0.10 for audio signal processing.

## 3. Results

### 3.1. Marginal Differences Between Base and Fine-Tuned Models

A characterization of classification performance across all aggregation statistics did not demonstrate a substantial advantage of fine-tuned models over the base wav2vec 2.0 model (Fig. 3). On average, the differences were negligible in terms of balanced accuracy: w2v2-base (0.76 ± 0.06) vs. w2v2-tuned-eng (0.76 ± 0.06) vs. w2v2-tuned-dysarthria (0.75 ± 0.06). Similarly small discrepancies were observed for the F1-score: w2v2-base (0.69 ± 0.10) vs. w2v2-tuned-eng (0.70 ± 0.09) vs. w2v2-tuned-dysarthria (0.70 ± 0.08). ROC-AUC values followed the same trend, with w2v2-base achieving 0.84 ± 0.06, compared to 0.83 ± 0.05 for w2v2-tuned-eng and 0.81 ± 0.05 for w2v2-tuned-dysarthria.

**Fig. 3.**
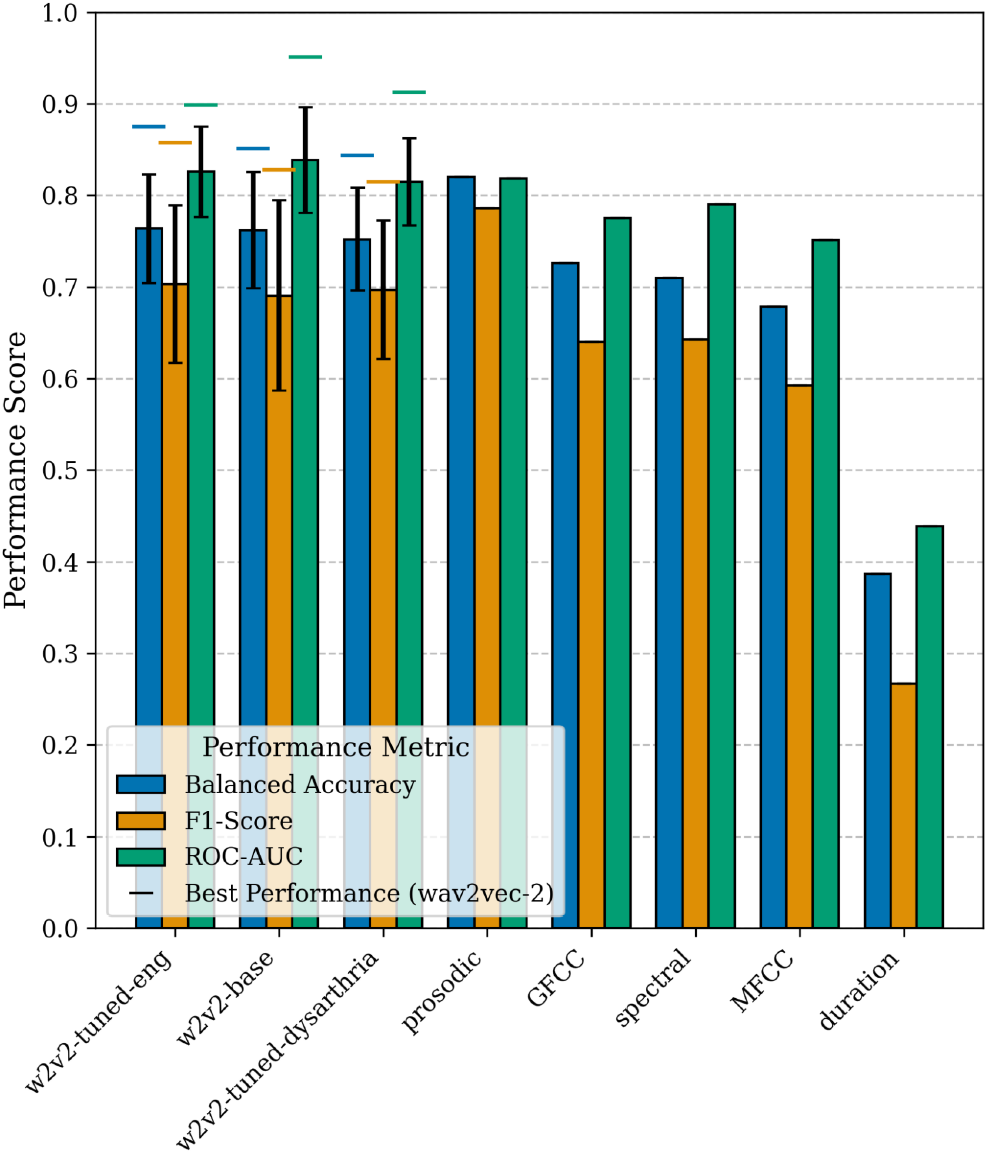
Classification performance for three wav2vec 2.0 variants: w2v2-base (layers 0, 1, 6, 11), w2v2-tuned-eng (layers 0, 1, 12, 23), and w2v2-tuned-dysarthria (layers 0, 1, 12, 23). All statistical aggregation functions were included for each bar (i.e. iqr, kurtosis, max, mean, median, min, q25, q75, skewness, std). Metrics are reported as mean ± standard deviation. Horizontal lines indicate the best-performing configuration within each wav2vec2 model. While averaging across all aggregation functions may dilute optimal performance, selected layer–aggregation combinations outperformed the baseline acoustic features.

When focusing on the best trained models, more variation was evident on the level of individual layers and aggregation functions. For w2v2-base, the highest balanced accuracy (0.85) and F1-score (0.83) occurred at the first Transformer layer using mean aggregation, whereas the best ROC-AUC (0.95) was obtained from layer 6 with q75 aggregation. In w2v2-tuned-eng, the best balanced accuracy (0.88) and F1-score (0.86) was observed at layer 0 with skewness aggregation, while ROC-AUC peaked at 0.90 for layer 0 with max or kurtosis aggregation. For w2v2-tuned-dysarthria, the top balanced accuracy (0.84) and F1-score (0.82) were achieved at layer 0 with min aggregation, while ROC-AUC reached 0.91 at layer 0 with skewness aggregation.

Among the baseline acoustic features, prosodic descriptors emerged as the strongest predictors (BA = 0.82, F1-score = 0.79, ROC-AUC = 0.82). Concatenating all acoustic feature sets slightly improved balanced accuracy and F1-score (BA = 0.84, F1-score = 0.82) but at the cost of reduced ROC-AUC (0.78). In contrast, naïve duration-based baselines failed to provide meaningful discrimination (BA = 0.39, F1-score = 0.27, ROC-AUC = 0.44).

### 3.2. Mean Aggregation Among the Top Strategies

Across the ten tested aggregation statistics (q25, median, mean, q75, std, kurtosis, min, iqr, skewness, max), mean pooling consistently ranked among the top three (q25, median, mean) and proved one of the most stable choices (Fig. 4, left). In particular, mean aggregation produced high median balanced-accuracy values and relatively small standard deviations for the w2v2-base model. The average balanced accuracy for the base model with mean pooling was BA = 0.81. Although mean pooling was robust overall, variability across aggregation functions became more apparent when comparing different model variants. The fine-tuned models exhibited greater layer-dependent fluctuation than the base model, and selected aggregation combinations occasionally outperformed the base configuration. Conversely, aggregation functions that emphasize extremes (min, max, kurtosis) tended to show a larger spread and lower medians across many settings.

**Fig. 4.**
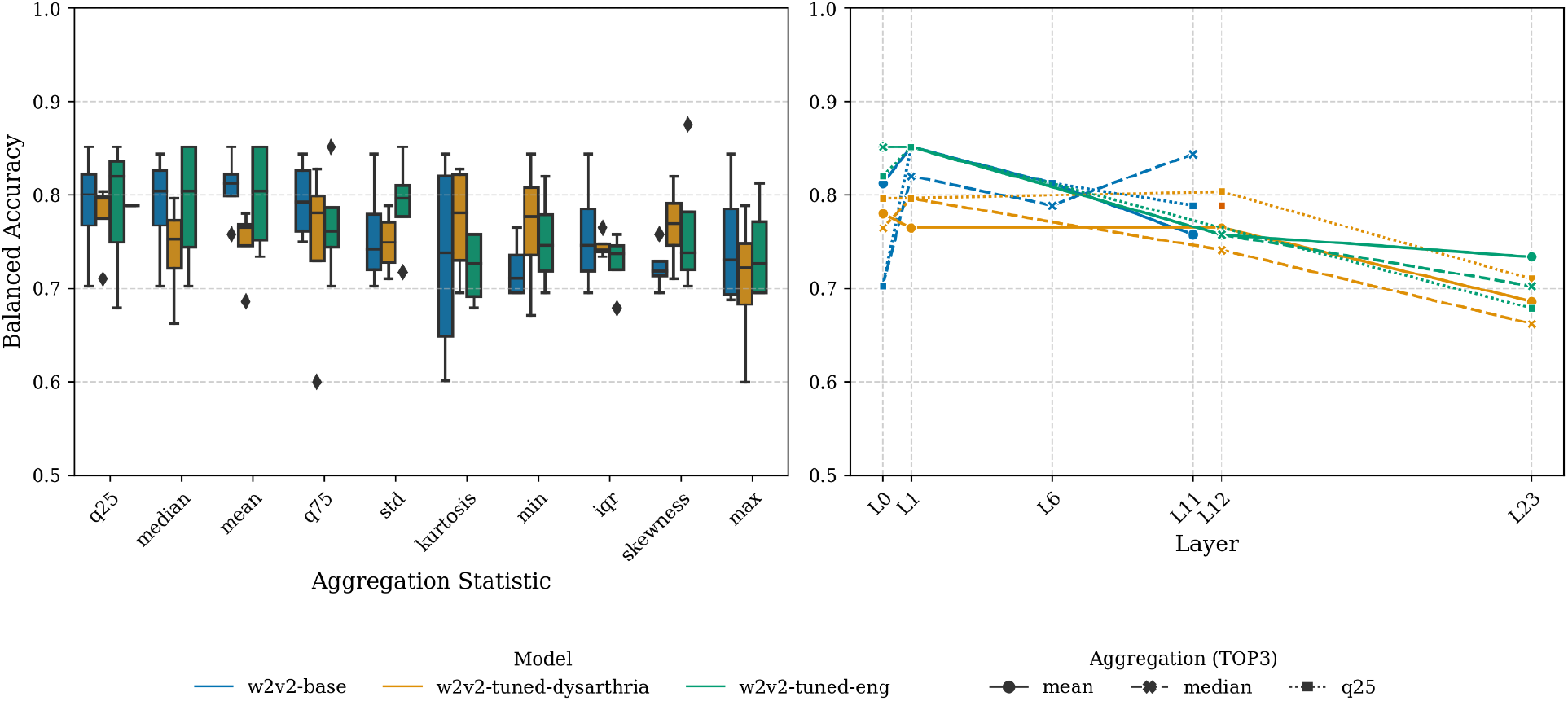
Balanced accuracy across aggregation functions and layer-wise trends. Left: Boxplots of balanced accuracy for ten aggregation statistics (q25, median, mean, q75, std, kurtosis, min, iqr, skewness, max) shown for three wav2vec 2.0 variants: w2v2-base (layers 0, 1, 6, 11), w2v2-tuned-eng (layers 0, 1, 12, 23), and w2v2-tuned-dysarthria (layers 0, 1, 12, 23). Right: Layer-wise balanced accuracy traces for the top three aggregations plotted at the layer indices (L0, L1, L6, L11/12, L23 as applicable).

Considering layer-wise trends (Fig. 4, right), the top three aggregations generally achieved their best balanced accuracy on early representations (L0–L1) for most models. However, exceptions were observed, for example, in the base model the median aggregation at a later layer (L11) produced performance comparable to that at layer L1.

### 3.3. No Added Benefit from Attention and Multi-Scale Pooling

Across the experiments with expanded layer coverage to include both early and deeper Transformer blocks, mean pooling consistently outperformed the more complex pooling strategies. Mean aggregation yielded the best and most stable results when evaluated on four metrics (balanced accuracy, F1 score, ROC-AUC, and precision) followed by multi-scale average pooling; attention-based pooling performed worst and showed substantially higher variance.

For the best strategy (mean pooling) the BA results were, w2v2-base: 0.80 ± 0.05; w2v2-tuned-eng: 0.80 ± 0.04; w2v2-tuned-dysarthria: 0.77 ± 0.03. The other scores followed a similar pattern (Fig. 5). Multi-scale pooling generally produced intermediate performance with lower variance than attention pooling, while attention pooling frequently suffered from large drops in ROC-AUC and precision for some fine-tuned variants.

**Fig. 5.**
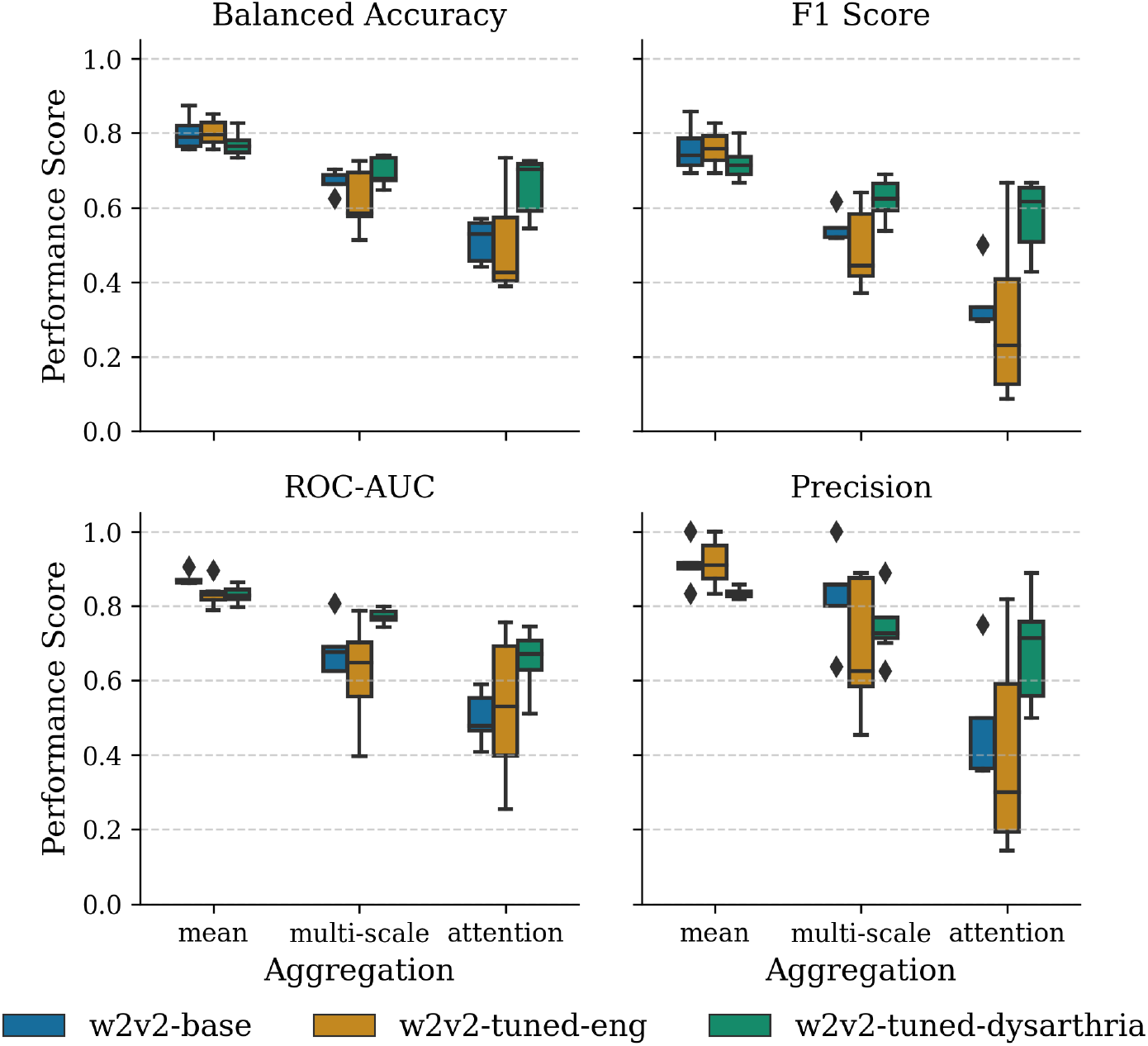
Comparison of aggregation strategies (mean, multi-scale, attention) across four performance metrics. Boxplots summarize cross-validation performance for three wav2vec 2.0 variants: w2v2-base (layers 1, 3, 6, 9, 11), w2v2-tuned-eng (layers 1, 4, 8, 12, 16, 20, 23), and w2v2-tuned-dysarthria (layers 1, 4, 8, 12, 16, 20, 23). Mean pooling was found to be the highest-performing strategy overall.

### 3.4. Feature Selection Enhanced Performance

This series of experiments focused primarily on the w2v2-base model and employed all ten statistical aggregation functions. The principal finding identified that the feature selection substantially improved classifier performance, with the ANOVA K-Best selector (SelectKBest) delivering the largest and most consistent gains (Fig. 6). K-Best stabilized performance across aggregation functions and Transformer layers. By contrast, PCA generally underperformed. It often degraded performance relative to using no selection, and performed substantially worse than K-Best in most settings.

**Fig. 6.**
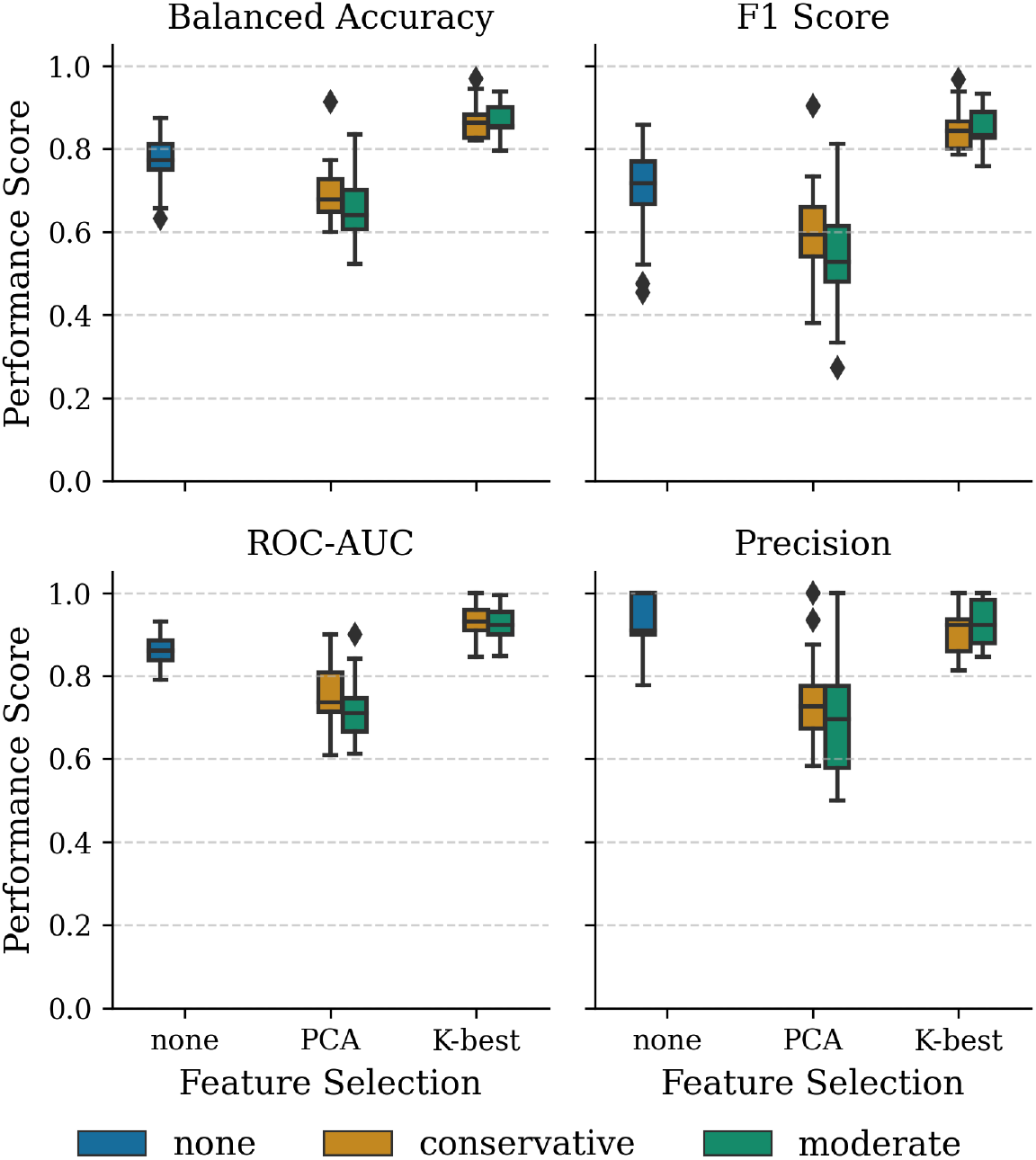
Effect of feature selection on classification performance for the w2v2-base model (layers 1, 3, 6, 9, 11). Boxplots show cross-validation distributions over all ten aggregation statistics and selected layers. Feature selection modes were: None (no selection), Conservative (top 12 features), and Moderate (top 20 features). K-Best (ANOVA F-value) stabilized and improved balanced accuracy.

In the conservative selection mode (12 features), the K-Best produced an average balanced accuracy of 0.87 and an average precision of 0.92 on the w2v2-base experiments. The best performing set of models using K-Best made only one to three false-negative errors (i.e., 1–3 PD subjects misclassified as healthy), with precision reaching 1.00. In those cases, the balanced accuracy exceeded 0.93, i.e., near-perfect discrimination on the given dataset. Specifically this was observed for mean aggregation at layer 6, q75 at layer 9, and mean at layer 11.

Feature selection on wav2vec-based embeddings yielded clear improvements, whereas applying feature selection to the traditional acoustic baseline did not produce similar benefits and even worsened the performance (Table 4).

**Table 4.** Acoustic-feature performance under different feature selection strategies.

| Feature set | Feature selection | Mode | Balanced Accuracy | ROC-AUC | Precision |
| --- | --- | --- | --- | --- | --- |
| all acoustic (211D) | None (211D) | - | 0.84 | 0.78 | 1.0 |
|  | PCA | moderate (20D) | 0.68 | 0.72 | 0.73 |
|  |  | conservative (12D) | 0.65 | 0.72 | 0.7 |
|  | K-best | moderate (20D) | 0.82 | 0.83 | 0.92 |
|  |  | conservative (12D) | 0.82 | 0.80 | 0.92 |

## 4. Discussion

### 4.1. Principal Findings

This study contributed to addressing the question of how frame-level wav2vec 2.0 embeddings should be aggregated into fixed-length utterance representations for the detection of PD from speech. The principal finding is that, although mean pooling was not universally superior across all models and layers, it consistently emerged as one of the most effective and stable aggregation strategies. These results support mean pooling as a simple, low-variance, and pragmatic default for wav2vec 2.0 representations in PD classification tasks. In addition, we proposed two innovative pooling schemes without introducing any trainable parameters, attention-based pooling and multi-scale average pooling, to test whether temporal weighting or multi-scale representations would improve robustness. Neither approach produced consistent benefits compared to statistical functions.

We also found that the differences between wav2vec 2.0 model variants were marginal. The base model performed comparably to fine-tuned alternatives, including one adapted on dysarthric corpora. This suggested that for English speech tasks, the wav2vec2-base model may suffice, without the need for specialized fine-tuning.

Interpreting the upper bound of obtained accuracy is essential. Prior research noted that PD symptom expression is heterogeneous across patients and presentations, which limits the generalizability of detection methods; dysarthria is estimated to occur in ∼90% of PD cases, not all [45, 46]. In our setting, a model achieving balanced accuracy ≈ 0.90 with precision = 1.0 can still miss ∼3–4 PD subjects in the MDVR-KCL corpus (i.e., false negatives) while making no false positives, i.e. consistent with high specificity (≈1.0) and a lower sensitivity (≈0.75–0.81), whose average yields BA ≈ 0.90. This aligns with the clinical reality that some PD speakers exhibit only subtle or no manifested acoustic impairment. In MDVR-KCL, 5/16 PD participants have UPDRS-II/III = 0, indicating no clinically rated speech/motor deficits. Thus, our error profile is coherent with known clinical heterogeneity (some PD cases may remain undetected by voice alone), while still showing that machine-learning models can capture subtle cues even when clinical ratings report no impairment.

### 4.2. Comparison With Prior Work

A growing number of studies have applied both handcrafted and learned representations to PD detection from speech using the MDVR-KCL corpus [47, 48, 49, 50, 51, 52, 53, 54]. Methods ranged from hybrid feature engineering plus classifiers to large self-supervised encoders. Many studies reported high discrimination on in-corpus evaluations (balanced accuracy and related metrics often close to or above 0.80–0.90), indicating that our findings are realistic, comparable, or even superior to top results in current literature.

Second, our layer-wise results, where early and mid layers often outperform the final Transformer outputs for PD detection, are consistent with recent analyses showing that the last hidden layer is frequently less useful for paralinguistic tasks. These findings align with evaluations reporting that intermediate layers typically retain more acoustic detail, whereas the top layer becomes overspecialized for ASR objectives; consequently, representations taken just before or from mid-depth Transformer blocks often yield superior downstream performance for pathology [55, 56] and emotion detection [57].

Importantly, building on the study “Aggregation Strategies of Wav2vec 2.0 Embeddings for Computational Paralinguistic Tasks” [29], and extending those ideas to dysarthric speech and PD, we concur with their conclusion that “mean aggregation is always a good choice, but it is not the only one.” In our experiments, a combination of mean aggregation, layer selection, and supervised feature selection appeared to be an effective pipeline for HC vs. PD discrimination.

### 4.3. Limitations and Future Directions

This study has certain limitations. The wav2vec variant fine-tuned on dysarthric corpora (TORGO, UASpeech) was developed for dysarthria, not specifically for PD. Moreover, the exact fine-tuning recipe and preprocessing available on the public model card are not fully documented. Consequently, conclusions about the value of dysarthria-adaptation for PD detection remain tentative and it is suggested that this should be confirmed using transparently reported fine-tuning procedures.

Our attention-based and multi-scale pooling implementations were deliberately non-parametric (no learnable weights). This design choice likely constrained their expressivity, i.e. without learned parameters the pooling schemes can only re-weight frames using fixed heuristics (L1 salience, fixed kernels), which may explain why they underperformed relative to simple statistical aggregation in our experiments. In other words, the poor performance of these non-learned pooling methods here does not preclude benefits from parameterized or trainable pooling mechanisms.

The study is limited to a single real dataset (MDVR-KCL) with a modest sample size. Although we made the research of practical importance, it may reduce the ability to generalize findings to other populations, recording conditions, and languages.

## 5. Conclusion

This paper showed that statistical pooling of frame-level wav2vec 2.0 embeddings outperformed more sophisticated temporal aggregation methods, and in combination with wav2vec2-base and targeted feature selection, it offered an accurate system for Parkinson’s disease detection from English read speech. We did not find any evidence that wav2vec models fine-tuned on multilingual or dysarthric corpora provide superior performance. The findings empirically support the continued use of mean pooling as a viable strategy for temporal aggregation of latent features for PD wav2vec-based detection.

## Data Availability

The dataset used is available at the following URL: https://zenodo.org/records/2867216.
All data produced in the present study are available upon reasonable request to the authors.

## Acknowledgement

This study was supported by the project of the National Institute for Neurological Research (Programme EXCELES, ID Project No. LX22NPO5107), funded by the European Union, Next Generation EU.

